# Geospatial Analysis of Multilevel Socio-environmental Factors Impacting the *Campylobacter* Burden Among Infants in Rural Eastern Ethiopia: A One Health Perspective

**DOI:** 10.1101/2024.07.03.24309853

**Authors:** Xiaolong Li, Dehao Chen, Song Liang, Jemal Yousuf Hassen, Sarah L. McKune, Arie H. Havelaar, Jason K. Blackburn, the CAGED Research Team

## Abstract

Increasing attention has focused on health outcomes of *Campylobacter* infections among children under five years in low-resource settings. Recent evidence suggests colonization of *Campylobacter* species contributes to environmental enteric dysfunction, malnutrition, and growth faltering in young children. *Campylobacter* species are zoonotic, and factors from humans, animals, and the environment are involved in transmission. Few studies have assessed geospatial effects of environmental factors along with human and animal factors on *Campylobacter* infections. Here, we leveraged *Campylobacter* Genomics and Environmental Enteric Dysfunction (CAGED) project data to model multiple socio-environmental factors on *Campylobacter* burden among infants in Eastern Ethiopia. Stool samples from 106 infants were collected monthly from birth through the first year of life (December 2020 – June 2022). Genus-specific Taqman real-time PCR was performed to detect and quantify *Campylobacter* spp. and calculate cumulative *Campylobacter* burden for each child as the outcome variable. Thirteen regional environmental covariates describing topography, climate, vegetation, soil, and human population density were combined with household demographics, livelihoods/wealth, livestock ownership, and child-animal interactions as explanatory variables. We dichotomized all continuous outcome and explanatory variables and built logistic regression models for the first and second half of the infant’s first year of life. Infants being female, living in households with cattle, reported to have physical contact with animals, or reported to have mouthed soil or animal feces had increased odds of higher cumulative *Campylobacter* burden in Eastern Ethiopia. Future interventions should focus on infant-specific transmission pathways and create adequate separation of domestic animals from humans to prevent potential fecal exposures.

## Introduction

*Campylobacter* species are zoonotic bacteria, and the animal reservoirs include poultry, free-living birds, ruminants such as cattle and goats, as well as other warm-blooded animals ^1,2^. Common symptoms of *Campylobacter* infection include fever, diarrhea, and abdominal pain ^3^. In addition to symptomatic infections, asymptomatic *Campylobacter* cases are common, specifically among children and adults in low-and middle-income countries (LMICs) ^4,5^. Evidence from studies in low-resource settings has shown that colonization of *Campylobacter* species acquired at the early stage of child development potentially contributes to environmental enteric dysfunction (EED), malnutrition, and growth faltering in children ^4,6,7^, which draws increasing attention to the long-term health effects of *Campylobacter* infections among children under five years of age (CU5) in LMICs.

*Campylobacter jejuni* and *C. coli* have been reported as the two most common species that cause illness in humans ^3^. Traditional methods selectively isolating these two species have resulted in a large body of knowledge specific to their disease burden and clinical manifestations ^8^. However, several *Campylobacter* species other than *C. jejuni/coli* (non-*C. jejuni/coli* species), such as *C. concisus*, *C. lari*, *C. upsaliensis*, and *C. ureolyticus,* have been recognized as “emerging species” and have shown increasing clinical importance ^9^. An increasing proportion of non-*C. jejuni/coli* species, including a new species "*Candidatus Campylobacter infans*” (*C. infans*), were isolated from child stool samples collected in low-resource settings, and several studies have revealed their potential linkages with child stunting ^7,10–13^. This evidence shifts our attention towards the genus *Campylobacter* as a whole, rather than focusing solely on *C. jejuni/coli* in the context of child health.

The classic “F-diagram” (fluids, fields, flies, fingers, fomites, and food) from WHO indicates that adequate sanitation and hygiene can be effective in interrupting the fecal-oral transmission routes of diarrheic pathogens, including *Campylobacter*, in low-resource settings ^14,15^. Driven by this idea, water, sanitation, and hygiene (WaSH) interventions, including improved pit latrine, hand-washing stations, liquid soap, and point-of-use water chlorination, have been developed and implemented in LMICs ^16,17^. Substantial evidence suggests that adequate WaSH contributes to reducing the risk of childhood diarrheal disease ^18,19^. However, several recent large-scale randomized controlled trials have found that WaSH interventions did not significantly reduce the occurrence of diarrhea or other enteric infections ^20–22^. One possible explanation is that traditional WaSH interventions generally target routes of children’s exposure to human feces but fail to pay adequate attention to children’s exposure to animal feces ^15,23^. Risk factors for exposing children to animal feces might include livestock ownership, proximity to livestock, cohabitation of humans and animals, etc.

The first year of a child’s life is marked by remarkable developmental milestones, and the differentiation between the first and second six months brings about major changes in both biological risk and environmental interaction. In the initial six months, breastfeeding plays a crucial role in offering infants essential nutrients and immune protection against pathogens ^24^. However, as the infant transitions into the second six months, the introduction of solid foods broadens the child’s nutritional spectrum but also amplifies their exposure to contaminants and pathogens through the food pathway ^25^. Meanwhile, the infant becomes more mobile, gaining the ability to explore their surroundings independently. This newfound mobility empowers infants to explore their environment actively through touching and tasting ^26^, potentially increasing exposure to pathogens via the fecal-oral pathway. This developmental transition underscores the dynamic effects of age on infant’s biological risk and environmental interaction.

Moreover, other environmental factors could impact the non-food transmission route of *Campylobacter* to humans directly or via animal reservoirs indirectly ^27,28^. Studies conducted in European countries, the United States, and Canada found campylobacteriosis incidence to be correlated with climatic variables, including temperature and precipitation ^29–33^. In addition to weather and climate factors, Sanderson et al. also examined the potential impacts of hydrology and landscape features, like soil type and land use, on the rates of human *Campylobacter* cases in the UK ^34^. This study showed that an increased risk of *Campylobacter* infections was associated with periods of high surface-water flow and catchment areas with cattle/sheep grazing on stagnogley soils. Another similar study conducted in New Zealand linked the risk of infection to a high dairy cattle density ^35^. However, these kinds of studies that investigate the influence of environmental factors have been predominantly conducted in developed countries with a focus on *C. jejuni/coli*. There is a critical knowledge gap in environmental effects on *Campylobacter* infections in LMICs.

Although multiple factors are involved in the transmission pathways of *Campylobacter*, few studies have assessed the combined effects of environmental covariates with other relevant factors from different domains including humans and animals. Since environmental data often comprise spatial information such as land cover, precipitation, and topography, geospatial analysis is needed to address the inherent differences in data types between environmental factors and human/animal factors by integrating them within a spatial framework (e.g., spatial regression). This capability to analyze data at different levels, from individual households to broader environmental landscapes, enhances the depth of risk factor identification. Coupled with the One Health perspective, this kind of analysis will help unravel the complex interplay between human, animal, and environmental factors involved in *Campylobacter* transmissions among children in a low-resource setting.

The longitudinal study of the *Campylobacter* Genomics and Environmental Enteric Dysfunction (CAGED) project conducted in rural Eastern Ethiopia aimed to examine the association between *Campylobacter* infection, related reservoirs, and child health outcomes ^36^. It collected not only the child fecal samples but also environmental (e.g., soil and drinking water) and livestock samples. A household questionnaire included components on people (e.g., demographics, livelihoods and wealth), livestock, and the interactions between people, livestock, and the environment. Combined with the child and environmental samples, these data provide an opportunity to investigate the potential combined effects of these risk factors on *Campylobacter* infection in infants.

## Materials and Methods

### Study Design and Protocol

A detailed description of the CAGED study design and protocol can be found elsewhere ^36^. Briefly, a total of 106 infants from 10 kebeles (the smallest administrative unit) of Haramaya woreda, Eastern Hararghe Zone, Oromia, Ethiopia (Figure 1) were enrolled at birth and followed until approximately 13 months of age. Written informed consent was obtained from both parents of each participating child in the local language (Afan Oromo). Household information, including demographics, livelihoods and wealth, livestock ownership, and child health and nutrition status, was collected through household surveys at baseline and endline, along with short surveys conducted monthly. During the study period, child stool samples were collected monthly and tested for *Campylobacter* species. In addition, fecal samples from mothers and siblings of the enrolled children, feces from livestock (i.e., chicken, cattle, goat, and sheep), and environmental samples (soil and drinking water) were collected biannually. DNA extraction, genus-specific Taqman real-time PCR, and species-specific Sybr Green real-time PCR were performed afterwards to detect, quantify, and characterize *Campylobacter* spp. in these samples ^37^.

**Figure 1.**
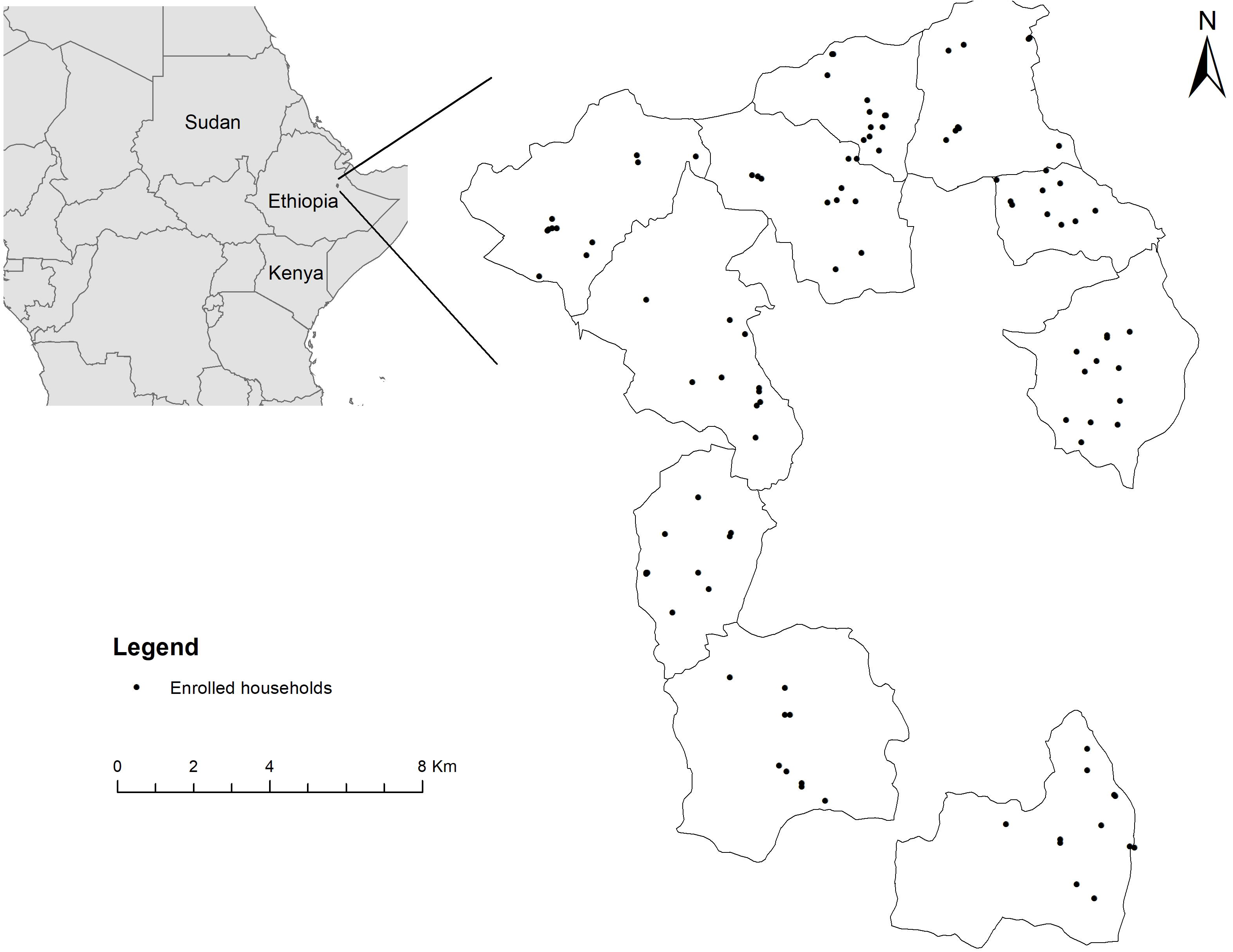
Study area and enrolled households in the *Campylobacter* Genomics and Environmental Enteric Dysfunction (CAGED) project.

### Outcome variable

We calculated the cumulative burden of *Campylobacter* infection for each enrolled child as the outcome variable of interest derived from the Ct values of their fecal samples collected over time. A genus-specific standard curve for expected bacterial load (log genome copies per 50 ng of DNA) against the Ct values was first generated using the 16S Taqman approach ^38^. One ml normalized bacterial culture cocktail (including *Campylobacter jejuni, Campylobacter coli, Campylobacter hyointestinalis, Campylobacter lari* and *Campylobacter fetus*) was used for DNA extraction, and 2 μl extracted DNA for qPCR afterwards. Nuclease free water and *Salmonella* genomic DNA were used as negative controls. We tested up to 10 *Campylobacter* DNA concentrations and repeated the experiment three times. Then the Ct values of the tested stool samples were converted to the expected bacterial load using this standard curve. The cumulative *Campylobacter* burden was defined as the average of the expected bacterial loads from available stool samples for each child ^37^.

### Explanatory Variables

#### Household surveys

Human and animal data were derived from household surveys. Demographic data including child sex, mother’s age, and mother’s education level were selected from the baseline survey, along with ownership of livestock (i.e., cattle, goat, sheep, and chicken) and assets (Table 1). To quantify all the livestock kept in a household, a composite metric, tropical livestock unit (TLU) was calculated based on the number of each species of livestock recorded in the baseline household survey ^39^.

**Table 1.** Demographics and socio-economic status of the households enrolled in this.

| Variable | N = 106<sup>\$</sup> |
| --- | --- |
| Sex |  |
| Female | 51 (48%) |
| Male | 55 (52%) |
| Mother's age | 27.0 (22.0, 32.0) |
| Unknown | 1 |
| Mother's education |  |
| No primary education | 76 (72%) |
| Some primary education | 29 (28%) |
| Unknown | 1 |
| Livestock ownership |  |
| Cattle | 52 (49%) |
| Goat | 60 (57%) |
| Sheep | 48 (45%) |
| Chicken | 53 (50%) |
| Tropical livestock unit | 0.62 (0.20, 1.40) |

<sup>\$</sup>n (%); Median (IQR)

In the monthly short surveys, we selected variables that reflect interactions between the target child and animals or the environment (e.g., physical contact with animals, crawling in areas with animal droppings, and mouthing of soil or animal feces), diet and nutrition (e.g., consumption of animal source food [ASF]), feeding practices (e.g., pre-lacteal feeding, introduction of complementary foods, and consumption of any solid food in the past 24 h), and use of antibiotics (i.e., being treated with antibiotics in the past month) (Table 2). In addition, two composite variables, minimum dietary diversity (MDD) and household food insecurity access score (HFIAS), were generated according to their respective literature ^40,41^.

**Table 2.** Variables used for regression modeling and univariate analysis results. Variable First period Second period.

| Variable | First period |  | Second period |  |
| --- | --- | --- | --- | --- |
|  | OR (CI)<sup>&</sup> | *p* | OR (CI) | *p* |
| Child sex | 0.8 (0.4, 1.7) | 0.56 | 2.3 (1.1, 5.1) | **0.03** |
| Mother's age | 1.3 (0.6, 2.8) | 0.55 | 0.8 (0.4, 1.7) | 0.55 |
| Mother's education | 1.2 (0.5, 2.9) | 0.49 | 1.1 (0.4, 2.6) | 0.49 |
| Cattle ownership | 0.6 (0.3, 1.4) | **0.24\*** | 2.1 (1.0, 4.7) | **0.05** |
| Goat ownership | 1.4 (0.6, 3.0) | 0.43 | 0.9 (0.4, 1.9) | 0.70 |
| Sheep ownership | 0.6 (0.3, 1.4) | **0.24** | 0.5 (0.2, 1.2) | **0.12** |
| Chicken ownership | 0.8 (0.4, 1.7) | 0.56 | 1.5 (0.7, 3.2) | 0.33 |
| Tropical livestock unit | 1.1 (0.5, 2.3) | 0.85 | 1.3 (0.6, 2.7) | 0.56 |
| Asset | 1.3 (0.6, 3.0) | 0.44 | 0.8 (0.4, 1.8) | 0.70 |
| Pre-lacteal feeding | 2.3 (1.0, 5.5) | **0.06** | NA | NA |
| Time of introduction of complementary foods (days) | 0.6 (0.3, 1.3) | **0.17** | NA | NA |
| Drinking from a bottle with nipple in the past 24h | 0.6 (0.3, 1.3) | **0.17** | 1.0 (0.5, 2.1) | 1 |
| Consumption of unpasteurized animal milk in the past 24h | 1.0 (0.2, 4.4) | 1 | 1.4 (0.6, 2.9) | 0.44 |
| Consumption of any solid food in the past 24h | 4.5 (1.1, 31.1) | **0.04** | 1.8 (0.9, 4.0) | **0.12** |
| Consumption of animal source food in the past 24h<sup>\$</sup> | NA | NA | 0.9 (0.4, 1.9) | 0.70 |
| Minimum dietary diversity<sup>\$</sup> | NA | NA | 0.8 (0.4, 1.7) | 0.56 |
| Household food insecurity access score | 1.3 (0.6, 2.7) | 0.56 | 2.3 (1.1, 5.1) | **0.03** |
| Vitamin-A supplementation in the past month<sup>#</sup> | NA | NA | 1.2 (0.5, 2.6) | 0.69 |
| Being treated with antibiotics in the past month | 1.6 (0.7, 3.7) | **0.22** | 0.8 (0.4, 1.7) | 0.55 |
| Contact with animals | 3.2 (1.4, 7.7) | **0.01** | 1.3 (0.6, 2.7) | 0.56 |
| Crawling in areas with animal feces | 3.0 (0.8, 14.2) | **0.11** | 1.5 (0.7, 3.2) | 0.33 |
| Mouthing of soil or animal feces | 12.1 (2.2, 226) | **0.00** | 1.5 (0.7, 3.2) | 0.33 |
| Preventive actions (mother) for mouthing behavior | 1.1 (0.5, 2.4) | 0.77 | 0.9 (0.4, 2.0) | 0.84 |
| Elevation (m) | 0.6 (0.3, 1.3) | **0.17** | 0.7 (0.3, 1.5) | 0.33 |

Table 2. Continued
| Variable | First period |  | Second period |  |
| --- | --- | --- | --- | --- |
|  | OR (CI) | *p* | OR (CI) | *p* |
| Maximum daily land surface temperature, 2021 (°C) | 1.3 (0.6, 2.7) | 0.56 | 1.1 (0.5, 2.3) | 0.85 |
| Mean daily land surface temperature, 2021 (°C) | 0.9 (0.4, 2.0) | 0.85 | 0.8 (0.4, 1.7) | 0.56 |
| Minimum daily land surface temperature, 2021 (°C) | 1.3 (0.6, 2.7) | 0.56 | 1.5 (0.7, 3.2) | 0.33 |
| Maximum 16-day NDVI<sup>‡</sup>, 2021 | 1.5 (0.7, 3.2) | 0.33 | 0.9 (0.4, 2.0) | 0.85 |
| Mean 16-day NDVI, 2021 | 1.3 (0.6, 2.7) | 0.56 | 0.7 (0.3, 1.5) | 0.33 |
| Minimum 16-day NDVI, 2021 | 0.8 (0.4, 1.7) | 0.56 | 0.5 (0.2, 1.1) | **0.08** |
| Population count 100m x 100m, 2020 | 1.2 (0.5, 2.5) | 0.70 | 0.3 (0.1, 0.6) | **0.00** |
| Slope (degree) | 0.4 (0.2, 0.9) | **0.03** | 1.3 (0.6, 2.7) | 0.56 |
| Proportion of clay particles in the fine earth fraction (g/kg) | 1.3 (0.6, 2.7) | 0.56 | 2.3 (1.1 to 5.1) | **0.03** |
| Soil organic carbon content in the fine earth fraction (0.1g/kg) | 0.9 (0.4, 1.8) | 0.70 | 1.2 (0.5, 2.5) | 0.70 |
| Soil pH | 0.5 (0.2, 1.3) | **0.16** | 0.8 (0.3, 2.0) | 0.64 |

& Odds ratio (confidence interval)
\* Bold script represents the *p*-value is less than 0.25, and the corresponding variable was included as a candidate for the multivariate analysis.
\$ Data were not available for the first period as the minimum dietary diversity (MDD) survey was administered after 6 months of age.

#### Environmental variables

Thirteen environmental covariates used for ecological niche modeling of the genus *Campylobacter* in a previous analysis ^42^ were also included in this study (Table 3). Corresponding values in grids within which the 106 enrolled households were located were extracted from the raster layer of each environmental covariate using the *raster* package ^43^ in R.

**Table 3.** Environmental variables used in this study and their sources. Variable Median Data source.

| Variable | Median | Data source |
| --- | --- | --- |
| Elevation (m) | 2083.0 | WorldPop (https://www.worldpop.org/) |
| Maximum daily land surface temperature, 2021 (°C) | 44.18 | MODIS Land Surface Temperature /Emissivity Daily (MOD11A1) Version 6.1 |
| Mean daily land surface temperature, 2021 (°C) | 31.17 | MODIS Land Surface Temperature /Emissivity Daily (MOD11A1) Version 6.1 |
| Minimum daily land surface temperature, 2021 (°C) | 16.58 | MODIS Land Surface Temperature /Emissivity Daily (MOD11A1) Version 6.1 |
| Maximum 16-day NDVI*, 2021 | 0.68 | MODIS Vegetation Indices (MOD13Q1) Version 6.1 |
| Mean 16-day NDVI, 2021 | 0.48 | MODIS Vegetation Indices (MOD13Q1) Version 6.1 |
| Minimum 16-day NDVI, 2021 | 0.30 | MODIS Vegetation Indices (MOD13Q1) Version 6.1 |
| Population count 100m x 100m, 2020 | 6 | WorldPop (https://www.worldpop.org/) |
| Slope (degree) | 5.0 | WorldPop (https://www.worldpop.org/) |
| Proportion of clay particles in the fine earth fraction (g/kg) | 421.5 | SoilGrids version 2.0 (https://soilgrids.org/) |
| Soil organic carbon content in the fine earth fraction (0.1g/kg) | 328 | SoilGrids version 2.0 (https://soilgrids.org/) |
| Soil pH | 7.4 | SoilGrids version 2.0 (https://soilgrids.org/) |

\* Normalized difference vegetation index

### Statistical Analysis

Given that feeding practices (exclusive breastfeeding vs. complementary feeding) and the motor ability of infants are most often quite different between the first and second half of infants’ first year of life, related risk factors such as probability of exposure to livestock and contaminated environments are likely to dynamically change over time. Child age is also an important confounding factor for *Campylobacter* infections ^4,39^. To be consistent with our previous analysis about the prevalence of *Campylobacter* by age groups ^42^, we split the whole study period into two parts, with a cut-off of 177 days of child age, which reflects the boundary between the first two and second two age groups classified in that analysis.

The outcome variable, cumulative *Campylobacter* burden, and all explanatory variables derived from the short household surveys were calculated separately for the two periods. We took the average of the monthly short survey data for each selected short survey variable, which resulted in an average value for the numeric variables or a proportion of being “1” for the binary variables (ordinal variable MDD was dichotomized using a generally accepted cut-off of < 5 and ≥ 5) during each period. Given the relatively small sample size, a median split approach was applied to dichotomize all continuous variables to improve the model robustness following a previous study ^44^.

The purposeful selection of covariates approach was employed in this study to choose the candidate variables and determine which to include in the final model ^45,46^. The likelihood ratio test from logistic regression was performed for each explanatory variable with the outcome variable. The univariate analysis was conducted for the two time periods separately with the same pool of explanatory variables. For the first period, two variables, consumption of ASF in the past 24 h and MDD, were excluded from the univariate analysis, as these questions were not asked until a child reached six months of age. Given 105 of 106 children didn’t have vitamin-A supplements in the first period, the variable vitamin-A supplementation was also excluded from the first period univariate analysis. Any variables with a *p*-value less than 0.25 were selected as candidates for the following multivariate analysis.

A multivariate logistic regression model was built first with all candidate variables, followed by an iterative variable selection process. In each iteration, the covariate with the maximum *p*-value in the model was identified first. If the covariate was not significant at the 0.1 alpha level, it was removed from the model temporally, and a reduced model was fit to compare the change of coefficients (Δβ) without that covariate. If the maximum Δβ was greater than 20%, which indicates a potential confounding effect of the excluded covariate with one or more of the remaining variables in the model, the excluded covariate was added back to the model, and the next covariable with the maximum *p*-value was evaluated. Otherwise, this temporarily excluded covariate was deleted from the pool of candidates, and a new multivariate model was built to start another round of evaluations. After multiple iterations, the final model with significant covariates and potential confounders was obtained.

Nagelkerke R^2^ ^47^, a so-called pseudo-R^2^ measure, and Hosmer & Lemeshow goodness-of-fit test ^48^ were used to test the model fit. For a sample size less than 200 and percentage of success in the outcome variable ranging from 38 – 62%, benchmark values between 0.32 and 0.58 of Nagelkerke R^2^ indicate good fit of the model ^49^. The Hosmer & Lemeshow test is a hypothesis test and evaluates if the expected event frequencies from the logistic regression model match the observed event frequencies in subgroups. The area under the receiver operating characteristic (ROC) curve (AUC) was used to assess the model performance in discriminating the positive results from the negatives ^50^. AUC values range from 0 to 1, and, empirically, values between 0.7 and 0.9 are a sign of good predictive performance. Values greater than 0.9 represent an excellent performance ^51^.

We first fitted a multivariate model using the data collected from the first period and tested if it was a good fit for the second period data. If not, a separate model was built for the second period, and effects of the covariates between two periods were evaluated. All the statistical analyses were performed using R version 4.1.1 ^52^.

### Spatial Autocorrelation Test

Spatial autocorrelation refers to the correlation within variables across different spatial units ^53^. If the values of a particular variable in nearby locations tend to be similar, a positive spatial autocorrelation exists in this variable, while negative spatial autocorrelation occurs when the values of a variable are more dissimilar than expected with their spatial neighbors. If spatial autocorrelation exists in the residuals of a regression model, it violates the assumption of independent errors and may result in an underestimation of the standard errors of the coefficient estimates of the model ^54^. To test the spatial autocorrelation in the residuals of the logistic models fit in this study, we performed Moran’s I test on the residuals using *spdep* package in R ^55^.

## Results

### Demographics and Socio-economic Status

Among the 106 enrolled children, 51.9% were male and 48.1% were female (Table 1). The mother’s age at baseline ranged from 17 to 43 years, with a median age of 27 years. A high proportion of mothers reported not attending school at any level; only 28% had some primary education. For livestock ownership, 49%, 57%, 45%, and 50% of households owned cattle, goat, sheep, and chicken, respectively. Accordingly, the indicator of tropical livestock units (TLU) kept by the household ranged from 0 to 5.54, with a median value of 0.62.

### Cumulative *Campylobacter* Burden

The calculated cumulative *Campylobacter* burden for infants in the first period ranged from 0.77 to 3.75 log genome copies per 50 ng of DNA with a median value of 2.10, while the minimum and maximum of cumulative *Campylobacter* burden for the second period were 1.95 and 5.06 log genome copies per 50 ng of DNA, respectively. The *Campylobacter* burden in the second half year of life (*M* = 3.52, *SD* = 0.75) was significantly higher than in the first half year of life (*M* = 2.14, *SD* = 0.62); *t*(210) = 14.7, *p* < .01 (Figure 2).

**Figure 2.**
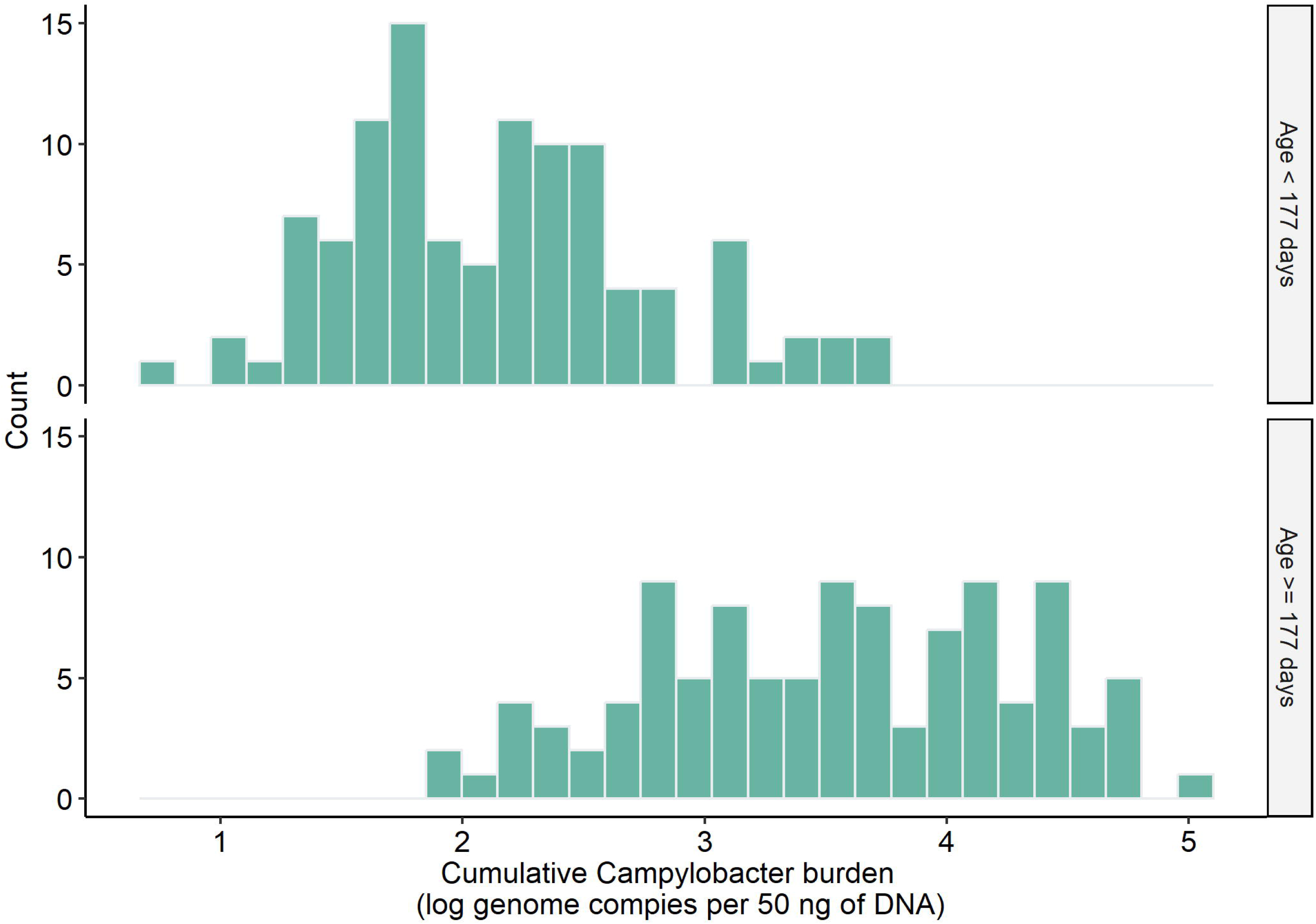
Frequency distribution of the cumulative *Campylobacter* burden for the first and second period in this study.

### Univariate Analysis

Based on the *p*-value of likelihood ratio test with a cut-off of 0.25, 13 and 8 variables were selected as candidate variables for the first and second period, respectively (Table 2). Among the candidates, three variables, namely, cattle ownership, sheep ownership, and consumption of any solid food in the past 24h, were included in the multivariate analysis for both periods. Though child sex was not significant at the alpha level of 0.25 in the first period, we still included it in the multivariate analysis given the confounding effect of child sex in *Campylobacter* infection reported in previous studies ^4,7,39,56^.

### Multivariate Analysis

#### Period One

The final logistic regression model for the first period showed that contact with animals, mouthing of soil or animal feces, and drinking from a bottle with nipple in the past 24h were statistically significant at the 0.05 level (Table 4). The direction of the estimated coefficients indicated children in the first period who had more physical contact with animals and more mouthing of soil or animal feces had greater odds of high cumulative *Campylobacter* burden, while drinking from a bottle with nipple was found to be a protective factor for high *Campylobacter* burden. Sheep ownership was marginally significant in the model and had a negative coefficient estimate. Through the purposeful selection of covariates process, child sex, pre-lacteal feeding, elevation, and soil pH were identified as potential confounding factors for *Campylobacter* burden.

**Table 4.** Logistic regression model for the cumulative *Campylobacter* burden in the first period.

| Characteristic | OR<sup>^</sup> | 95% CI<sup>*</sup> | *p*-value |
| --- | --- | --- | --- |
| Child sex | 0.59 | (0.22, 1051) | 0.3 |
| Sheep ownership | 0.40 | (0.15, 1.01) | 0.06 |
| Pre-lacteal feeding | 1.77 | (0.66, 4.87) | 0.3 |
| Contact with animals | 3.13 | (1.11, 9.6) | **0.04**<sup>\$</sup> |
| Mouthing of soil or animal feces | 12.8 | (1.80, 272) | **0.03** |
| Drinking from a bottle with nipple in the past 24h | 0.35 | (0.12, 0.94) | **0.04** |
| Elevation | 0.61 | (0.23, 1.60) | 0.30 |
| Soil pH | 0.43 | (0.14, 1.29) | 0.14 |
| Nagelkerke R<sup>2</sup> | 0.40 |  |  |
| Hosmer & Lemeshow test |  |  | 0.42 |
| AUC<sup>#</sup> | 0.79 | (0.70, 0.88) |  |
| Moran's I statistic | -0.06 |  | 0.75 |

<sup>^</sup>Odds ratio
<sup>\*</sup>Confidence interval
<sup>\$</sup>Boldness represents the *p*-value is less than 0.05.
<sup>#</sup>Area under the receiver operating characteristic curve

The Nagelkerke R^2^ of the logistic regression model was 0.40, which fell into the benchmark values (0.32 – 0.58) for good model fit (Table 4). Hosmer & Lemeshow test also showed there was no evidence of poor model fit. The model also had a relatively good predictive performance with an AUC of 0.79 (95% CI: 0.70 – 0.88) (Figure 3A). Moran’s I statistics showed that no spatial autocorrelation existed in the model residuals, suggesting there was no need to consider a spatial regression model to account for the spatial autocorrelation.

**Figure 3.**
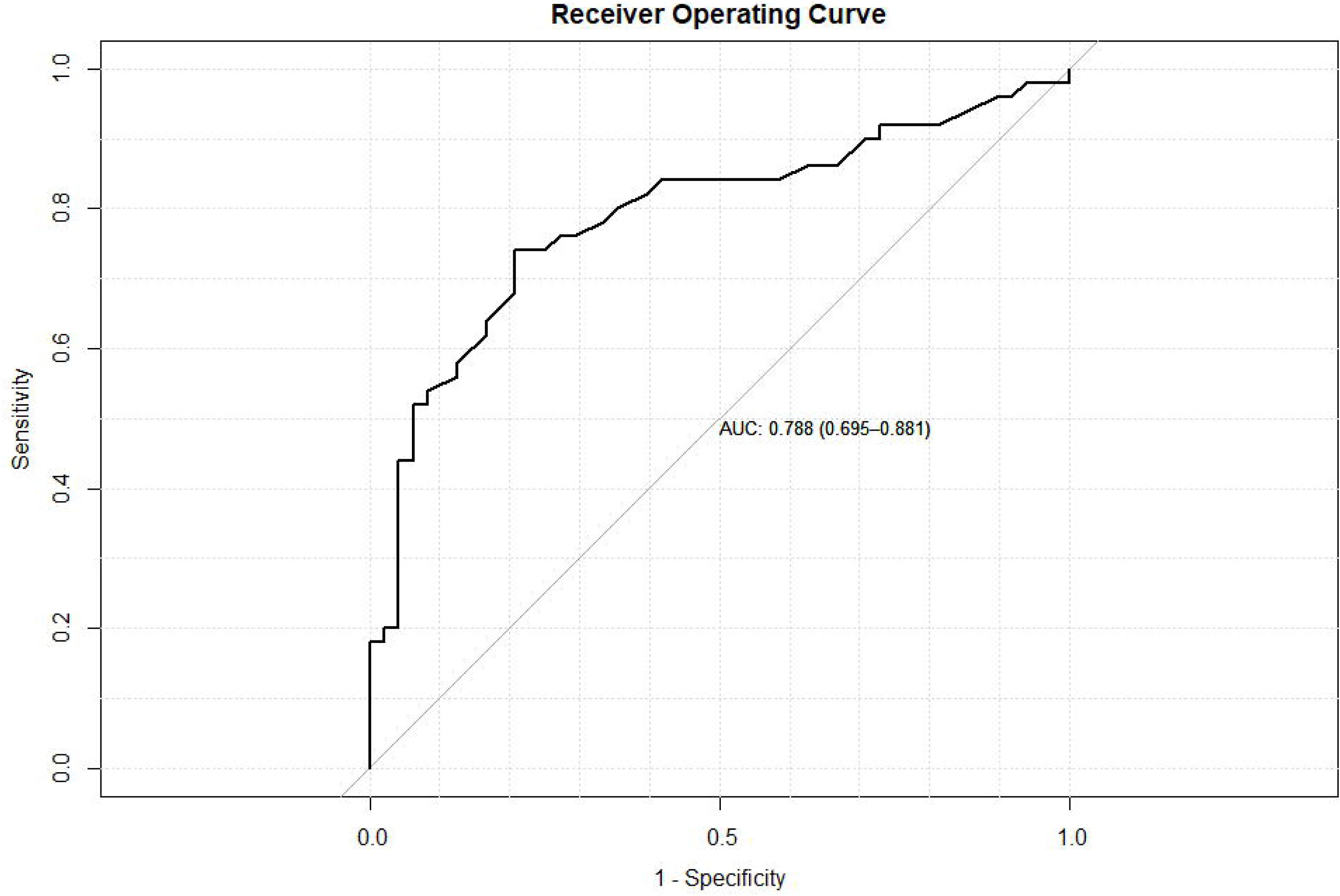
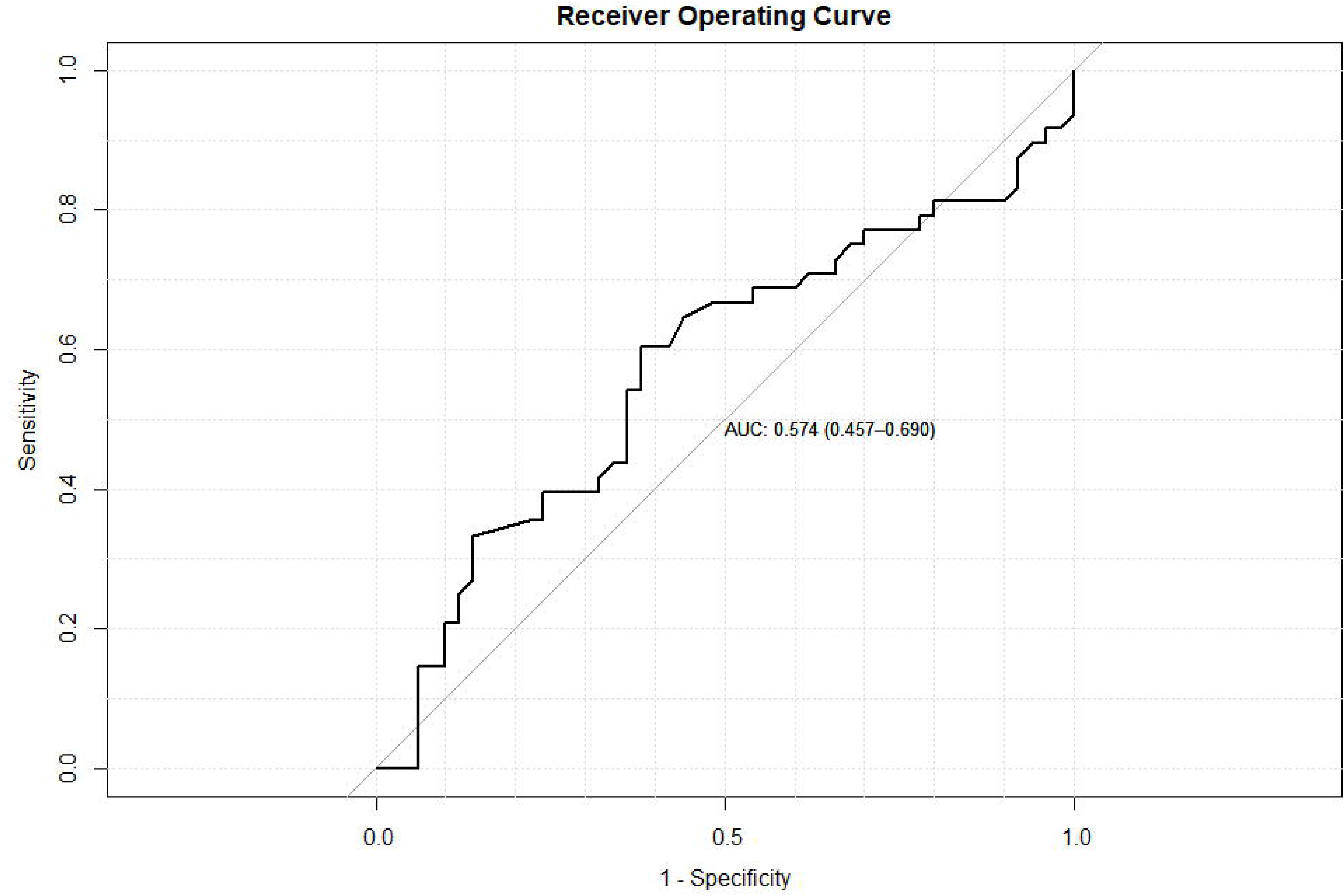
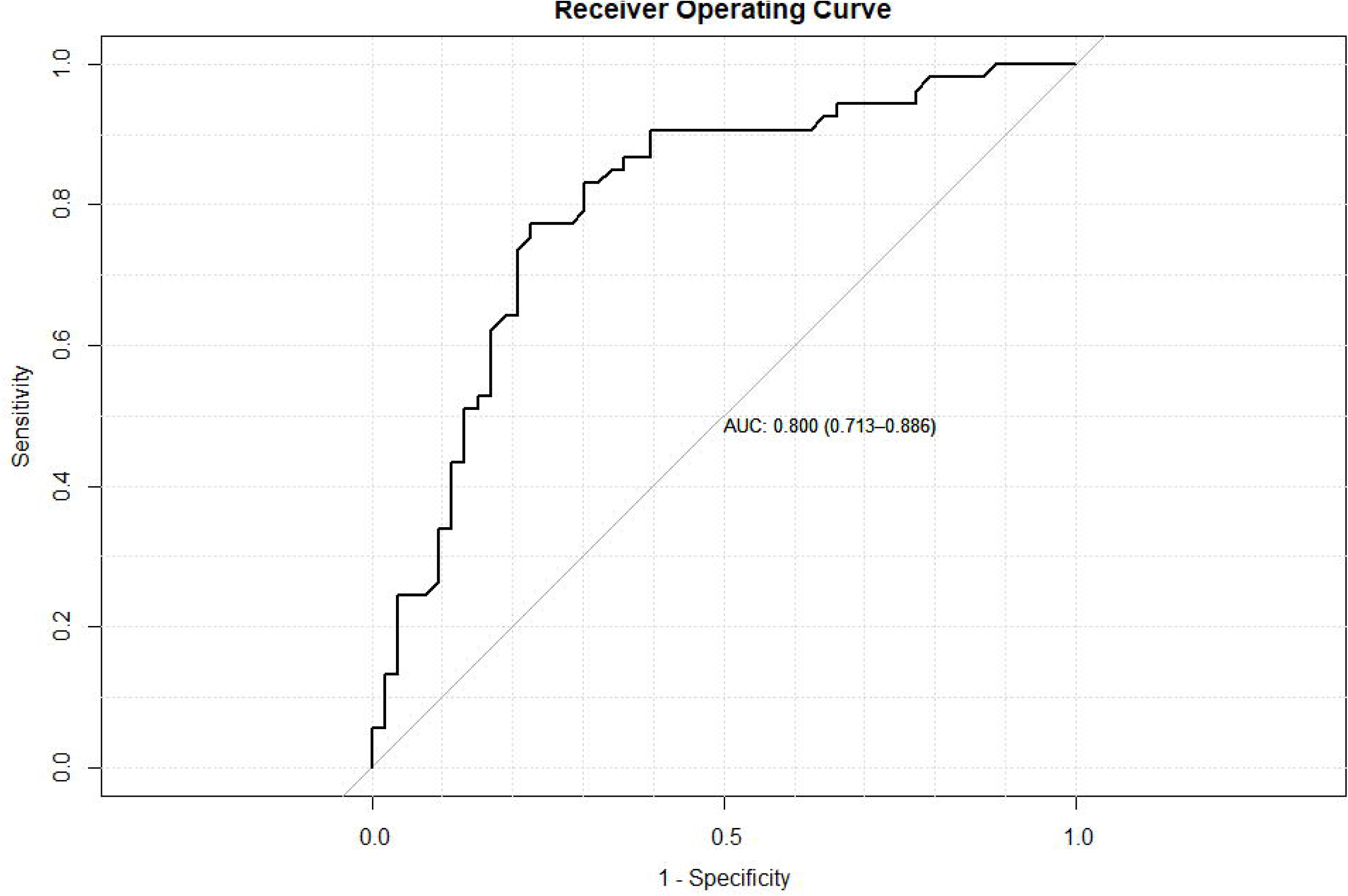
Receiver operating curves (ROC) for logistic regression models fit in this study. A: model fit with the first period data; B: test of the fit of the first period model to the second period data; C: model fit with the second period data.

To test if one single model can be obtained for both periods, the second period data were plugged into the model for the first period as a testing data set. Hosmer & Lemeshow test checked the difference between observed data and predicted values and showed this difference was significant (*p* = 0.00), suggesting a poor fit of the model to the second period data. It was also supported by a low AUC of 0.57 (95% CI: 0.46 – 0.69) (Figure 3B).

#### Period two

Logistic regression models were then fitted using the candidate variables selected from the univariate analysis for the second period, and the final model included eight variables with child sex, cattle ownership, and population density (per 100m^2^) being statistically significant at the 0.05 level (Table 5). Children being female and living in households that kept cattle had greater odds of high cumulative *Campylobacter* burden, while population density showed protective effects on *Campylobacter* burden. Household food insecurity access score, proportion of clay particles in soil, consumption of any solid food in the past 24h, and sheep ownership were identified as potential confounding factors for *Campylobacter* burden in the second period.

**Table 5.** Logistic regression model for the cumulative *Campylobacter* burden in the second period.

| Characteristic | OR<sup>^</sup> | 95% CI* | p-value |
| --- | --- | --- | --- |
| Child sex | 2.85 | (1.15, 7.47) | **0.027**<sup>\$</sup> |
| Cattle ownership | 2.87 | (1.14, 7.70) | **0.029** |
| Sheep ownership | 0.49 | (0.91, 1.21) | 0.12 |
| household food insecurity access score | 1.78 | (0.69, 4.59) | 0.2 |
| Consumption of any solid food in the past 24h | 2.01 | (0.79, 5.30) | 0.15 |
| Minimum 16-day NDVI, 2021 | 0.43 | (0.17, 1.05) | 0.069 |
| Population count 100m x 100m, 2020 | 0.37 | (0.15, 0.92) | **0.033** |
| Proportion of clay particles in the fine earth fraction | 2.12 | (0.84, 5.54) | 0.12 |
| Nagelkerke R<sup>2</sup> | 0.34 |  |  |
| Hosmer & Lemeshow test |  |  | 0.47 |
| AUC<sup>#</sup> | 0.80 | (0.71, 0.89) |  |
| Moran's I statistic | 0.014 |  | 0.37 |

<sup>^</sup>Odds ratio
<sup>\*</sup>Confidence interval
<sup>\$</sup>Bold script represents the p-value is less than 0.05.
<sup>#</sup>Area under the receiver operating characteristic curve

The Nagelkerke R^2^ for the second model was 0.34, and Hosmer & Lemeshow test showed there was no evidence of poor model fit (Table 5). This model had a good predictive performance with an AUC of 0.80 (95% CI: 0.71 – 0.89) (Figure 3C). Again, there was no spatial autocorrelation existing in the model residuals.

## Discussion

From a One Health perspective, this study identified potential factors involved in the *Campylobacter* transmission pathways between humans, animals, and the environment in a low-resource setting. Child age-specific behaviors and other factors that may increase the risk of children’s exposure to animal feces are largely missing from previous interventions that aimed to mitigate the burden of *Campylobacter* infection or other enteric illnesses among infants and young children in LMICs ^57^. Risk factors of *Campylobacter* infections that have been commonly identified in previous studies conducted among CU5 in low-resource settings are primarily limited to maternal educational level, feeding practices, WaSH indicators, and ownership of domestic animals ^4,58–60^. However, fewer studies have delved into infant-specific behaviors and the interactions between infants and livestock, which could also contribute to the risk of *Campylobacter* infections ^61^. Considering the dynamic effects of age, particularly distinguishing between infants aged less than six months and those aged six to twelve months, on biological risk and environmental interaction, our study introduces a novel approach. By fitting models and identifying risk factors separately for the first and second six months of infancy, we aim to capture the nuanced variations in infection dynamics during these critical developmental stages. This innovative methodology, coupled with the exploration of previously overlooked factors using longitudinal data, sets our study apart from existing research on *Campylobacter* infection risk factors.

At an early stage of life, infants and young children put non-food objects in their mouths as one way to explore the surrounding environment ^62^. However, this exploratory behavior could put children at higher risk of contracting zoonotic pathogens if they live in an environment contaminated by livestock and poultry feces. Our results identify mouthing of soil or animal feces as a significant risk factor contributing to a higher cumulative *Campylobacter* burden during the first half of the first year of life. This association can be attributed to the high prevalence of *Campylobacter* spp. in the feces of animals, including domestic livestock and poultry. Results from the laboratory showed that the prevalence of *Campylobacter* in the fecal samples of cattle, sheep, goats, and chicken collected from the CAGED longitudinal study were 99%, 98%, 99%, and 93%, respectively ^37^. In rural areas of LMICs, infants and young children are frequently placed on the ground sharing space with free-ranging livestock ^57^. As animals defecate in the homestead, or sometimes even inside the homes, enteric pathogens harbored in livestock feces or the soil contaminated by the feces may be ingested by infants through their routine mouthing behaviors while they are on the floor. Ingestion of soil and livestock feces is therefore a point of exposure specific to infant and young child behavior. This infant-specific transmission pathway needs to be considered in the future design of intervention strategies for preventing children from fecal exposure. This finding echoes the collective conclusion from the WASH Benefits and the Sanitation, Hygiene, Infant Nutrition Efficacy (SHINE) trials; that more effective interventions are needed to reduce the exposure to fecal contaminations in the domestic environments other than the traditional WaSH interventions ^63^.

Similarly, physical contact with animals was identified as another risk factor existing in the child-livestock interaction in the first period (before six months of age) that increases the exposure of children to animal feces. It is very common that people in rural areas share living and sleeping quarters with their livestock, which frequently exposes young children in the household to direct contact with livestock. Keeping animals inside human living spaces has been associated with *Campylobacter*-positive child stools in another study in rural Ethiopia ^57^, highlighting the need to design interventions creating adequate separation of domestic animals from the human living spaces to block the transmission pathway through direct contact.

The risks and benefits of raising livestock in smallholder families on child health are intertwined ^23^. On one hand, livestock raised in the household can provide animal source foods, which are seen as the best source of nutrition-rich food for infants and young children. Livestock production is also a source of household income. Increased income from livestock sales can grant families more purchasing power for food, which further helps improve child nutritional status. However, livestock ownership can potentially increase the risk of children’s exposure to animal feces, as discussed above. In this study, we didn’t find a significant association between consumption of animal source food and *Campylobacter* burden, but cattle ownership was identified as a significant factor associated with increased odds of having higher cumulative *Campylobacter* burden during the second half of the first year of life. In our study population, when cattle were kept either inside the homes or on the homestead but outside the home during the day, 100% (27/27) and 93.3% (28/30) of households, respectively, confined those cattle. During the night, almost all cattle were kept and tied inside the home. Regarding the purposes of raising cattle, households reported selling livestock for income (44.2%; 23/52) and consumption of milk (51.9%; 27/52) as the two major purposes. These data support other findings indicating a strong cultural focus on milk consumption in this area, which may correspond with an underlying risk in the consumption of raw/unpasteurized milk which has been associated with reported outbreaks of campylobacteriosis in HICs ^64,65^. A recent Ethiopian study showed a higher prevalence of *C. jejuni* (16%) in raw milk compared to other dairy products ^66^. Though consumption of raw milk was not identified in our models as a significant risk factor of *Campylobacter* burden in our study population, further work is still needed to unpack this association among children in low-resource settings.

Appropriate infant and young child feeding practices could improve child nutritional status, growth, and development ^67^. For infant aged from 0 – 5 months, exclusive breastfeeding is strongly recommended by WHO, while, bottle feeding, using a bottle with a nipple/teat to feed any liquid or semi-solid food, is discouraged at this early stage of life due to global concerns that include excessive weight gain, iron depletion, etc. ^68^. In addition, a bottle with a nipple is more likely to be contaminated in low-resource settings where inadequate cleaning and disinfection of bottles is more common, which increases the risk of enteric infections ^69^. Lengerh et al. showed that bottle feeding was associated with increased odds of *Campylobacter* infection among diarrheic children in northwest Ethiopia ^60^. However, in this study, drinking from a bottle with nipple was shown to be a protective factor for higher cumulative *Campylobacter* burden in the first time period. Our results should not be interpreted as an encouragement of bottle feeding. One potential explanation could be that this practice is linked to other socio-economic factors that contribute to reducing the risk of *Campylobacter* infections. A previous study using Ethiopian Demographic and Health Surveys data (EDHS) to examine the determinants of bottle feeding suggested that women who had a higher education, came from a richer household, and lived in urban areas were more likely to bottle feed ^68^. The role of mother’s education level and household wealth status in *Campylobacter* burden was not clear in our analysis with a relatively small sample size, and further work is needed to unpack these potential links.

Influences of environmental and climatic factors have been rarely reported in studies investigating risk factors of *Campylobacter* infections among children in LMICs. Here, we included several environmental covariates previously used to model the prevalence of enteric diseases globally and regionally. Due to the small geography of our study area, most environmental covariates have less heterogeneity to show signals in their association with *Campylobacter* infections. This is one of the limitations in this study. Evaluating these environmental effects on *Campylobacter* infections at a larger scale in LMICs would be a future direction.

We tested the spatial autocorrelation in the model residuals during the model-building process, a step that has seldom been taken in previous work on identifying risk factors of *Campylobacter* infections. We did so not only because we included environmental covariates in the models, but to consider the potential spatial effects (i.e., spatial dependence and spatial heterogeneity) introduced by georeferenced data ^70^. Neglecting the spatial effects could lead to an inflation of variance in regression estimates and, consequently, a less reliable regression model ^71^. Therefore, it would be appropriate for future studies to consider including the spatial autocorrelation test as a core component of the modeling-building process to ensure the reliability and accuracy of the regression model.

Another innovative aspect of this study involves the adoption of bacterial load-based cumulative burden, departing from traditional prevalence measures when dealing with longitudinal data. By calculating the cumulative burden, we gain insight into the persistent impact of *Campylobacter* infections over time, offering a nuanced perspective on disease dynamics. This method enables a comprehensive assessment of *Campylobacter* burden over a certain period and holds promise for advancing longitudinal studies on *Campylobacter* infections.

In conclusion, our study reveals that factors involved in the interactions between human (infants), livestock, and home environment impacted the presence of higher cumulative *Campylobacter* burden among infants in eastern Ethiopia. For young infants aged less than 6 months, being reported to have physical contact with animals and have mouthing of soil or animal feces were identified as risk factors of higher *Campylobacter* burden. And drinking from a bottle with nipple was shown to be a protective factor. This result requires additional research to understand and should not be interpreted to encourage bottle feeding; additional information is required to understand whether there is a direct causal mechanism or whether underlying factors or confounders, such as socio-economic status or overall household hygiene, might explain the finding. While older infants (aged between 6 and 12 months) being female and living in households with cattle had increased odds of higher *Campylobacter* burden. High population density (potentially linked to urban residency) was identified as a protective factor for this age group. Future interventions should pay more attention to the infant-specific transmission pathway and create adequate separation of domestic animals from humans to prevent infants and young children from potential fecal exposures.

## Data Availability

Deidentified individual participant data will be made available through Dataverse (https://dataverse.org/) after December 31, 2024.

## Acknowledgments

This work is a result of the CAGED Research Team whose members include: Amanda Evelyn Ojeda, Arie H. Havelaar, Abadir Jemal Seran, Abdulmuen Mohammed Ibrahim, Bahar Mummed Hassen, Belisa Usmael Ahmedo, Cyrus Saleem, Dehao Chen, Efrah Ali Yusuf, Gireesh Rajashekara, Getnet Yimer, Ibsa A. Ahmed, Ibsa Aliyi Usmane, Jafer Kedir Amin, Jason K. Blackburn, Jemal Y. Hassen, Kedir A. Hassen, Kunuza Adem Umer, Karah Mechlowitz, Kedir Teji Roba, Loic Deblais, Mussie Bhrane, Mark J. Manary, Mawardi M. Dawid, Mahammad Mahammad Usmail, Nigel P. French, Nur Shaikh, Nitya Singh, Sarah L. McKune, Wondwossen A. Gebreyes, Xiaolong Li, Yenenesh Demisie Weldesenbet, Yang Yang, Zelalem Hailu Mekuria.

## Financial Support

This project is funded by the United States Agency for International Development Bureau for Food Security under Agreement #AID-OAA-L-15-00003 as part of Feed the Future Innovation Lab for Livestock Systems, and by the Bill & Melinda Gates Foundation OPP#1175487. Under the grant conditions of the Foundation, a Creative Commons Attribution 4.0 Generic License has already been assigned to the Author Accepted Manuscript version that might arise from this submission. Any opinions, findings, conclusions or recommendations expressed here are those of the authors alone. Research reported in this publication was supported by the University of Florida Clinical and Translational Science Institute, which was supported in part by the NIH National Center for Advancing Translational Sciences under award number UL1TR001427.

## Authors contributions

XL, SL, SLM, AHH and JKB conceptualized and designed the study; XL and DC curated and analyzed data; JYH, SLM, and AHH managed and coordinated the research activity in this project; XL drafted the initial manuscript; and SL, DC. SLM, AHH, JKB reviewed and critically revised the manuscript. All authors approved the final manuscript as submitted and agreed to be accountable for all aspects of the work.

## Authors contact information

Xiaolong Li, Department of Environmental and Global Health, College of Public Health and Health Professions, University of Florida, Gainesville, FL; Dehao Chen, Department of Environmental and Global Health, College of Public Health and Health Professions, University of Florida, Gainesville, FL; Song Liang, Department of Environmental and Global Health, College of Public Health and Health Professions, University of Florida, Gainesville, FL; Jemal Yousuf Hassen, Haramaya University, Dire Dawa, Ethiopia; Sarah L. McKune, Department of Environmental and Global Health, College of Public Health and Health Professions, University of Florida, Gainesville, FL; Arie H. Havelaar, Department of Animal Sciences, Institute of Food and Agricultural Sciences, University of Florida, Gainesville, FL; Jason K. Blackburn, Spatial Epidemiology and Ecology Research Laboratory, Department of Geography, College of Liberal Arts and Sciences, University of Florida, Gainesville, FL.

## Notes

### Competing Interest Statement

The authors have declared no competing interest.

### Author Declarations

Internal Review Board of the University of Florida (IRB201903141); the Institutional Health Research Ethics Committee at Haramaya University (COHMS/1010/3796/20), and the Ethiopia National Research Ethics Review Committee (SM/14.1/1059/20) each gave approval for this work.

